# Co-coverage of social protection programs and maternal and child nutrition interventions in Bangladesh, India and Nepal

**DOI:** 10.1101/2025.01.22.25320966

**Authors:** Samuel Scott, Sumanta Neupane, Hana Tasic, Nadia Akseer, Sunny S. Kim, Harold Alderman, Bianca Carducci, Rebecca Heidkamp

## Abstract

Social protection programs (SPPs) are common in South Asia, a global malnutrition hotspot. Provision of SPP benefits as well as essential health/nutrition interventions to mothers and children are goals for optimal health and development outcomes, but the degree of co-coverage of SPPs and health/nutrition interventions among beneficiary households is poorly described. Using six population-based surveys from 2012 to 2019 in Bangladesh, India, and Nepal (n=253,703 women with children under five years of age), we examined data availability for SPPs (food and cash transfers) and health/nutrition interventions, and estimated their coverage and co-coverage during a woman’s last pregnancy (interventions: take-home food rations plus at least four antenatal care visits, receipt of at least 100 iron-folic acid tablets, deworming, and tetanus injections), after delivery (cash benefit plus the interventions above), and in children (take-home ration for the child plus vitamin A supplementation, deworming, iron syrup, growth monitoring, and nutrition counseling). In India, 52% and 51% of women and children, respectively, received food transfers, but only 3% and 8% received food plus all health/nutrition interventions. In India and Nepal, respectively, cash after delivery was received by 41% and 86% of women, but only 2% and 21% received cash after delivery plus all health/nutrition interventions. There was insufficient data to estimate coverage of both SPPs and health/nutrition interventions in Bangladesh. Our findings highlight the need for data on both SPP and health/nutrition intervention coverage in household surveys. There are missed opportunities to reach women and children with interventions across multiple sectors.

## INTRODUCTION

Forty percent of households (809 million individuals) in South Asia experienced moderate or severe food insecurity in 2022, and 30% of children under five years old in the region are stunted (1). It is well-known that nutrition interventions alone will not eradicate poor nutrition outcomes; solutions from multiple sectors that target underlying and immediate determinants of these outcomes are needed (2,3). Cash or in-kind transfers offered through social protection programs (SPPs), while not necessarily designed with nutrition outcomes in mind, can address underlying determinants missed by nutrition interventions and, in some cases, have been effective at reducing undernutrition in South Asian contexts (4). Making SPPs more “nutrition-sensitive” by adding features that may enhance program impacts on nutrition outcomes has been recommended (5,6). For example, a cash transfer may provide relief to a household’s budget but, without an additional intervention such as information on the importance of consuming nutritious foods, the transfer may not lead to impacts on diet and nutrition, as was observed in a trial in Bangladesh (7). A recent review of SPPs in South Asia suggests that there is ample scope for making existing programs more nutrition-sensitive through targeting vulnerable households, including nutrition behavior change, linking the program to health services, empowering women, providing or ensuring access to foods with a high nutritional value, or including other measures to promote healthy diets (8).

While it is important to track the reach of SPPs over time, large-scale scale household surveys have limited data on exposure to SPPs and their potentially nutrition-sensitive features (9). Thus, assessing coverage of nutrition-sensitive SPPs is currently hindered by poor data availability and measurement challenges. To understand if households are reached simultaneously by multiple interventions, estimated indicators of co-coverage (i.e., the percentage of households receiving more than one intervention of interest) are an alternative method of analysis. Co-coverage has been used to understand simultaneous coverage of multiple health/nutrition interventions within the same household (10–12). A recent non-representative phone survey in India found low co-coverage of food and cash transfers with health/nutrition interventions among mothers (13). In the current study, using data from representative population-based surveys that measure household exposure to SPPs, we estimate co-coverage of SPPs with health/nutrition interventions during pregnancy and childhood in Bangladesh, India, and Nepal.

## METHODS

### Data sources and classification of SPPs

The following publicly available, nationally representative surveys with data on SPPs were included: 1) Bangladesh Household Income and Expenditure Survey (HIES) 2016; 2) Bangladesh Multiple Indicator Cluster Survey (BMICS) 2019; 3) India National Family Health Survey (NFHS) 2016; 4) India National Sample Survey (NSS) 2012; 5) Nepal Demographic and Health Survey (NDHS) 2016; 6) Nepal Multiple Indicator Cluster Survey (NMICS) 2019. While population-based surveys in other South Asian countries exist, they were not included in this study due to being older than 10 years, limited accessibility, or not having data on SPPs. The World Bank ASPIRE framework (14) was used to organize the SPPs by five categories of cash for work, unconditional cash transfers (poverty-targeted cash, allowances, and public charity), conditional cash transfers, food transfers, and school feeding. In these six surveys, data from women with children under five years old were used. All women provided consent for participation in the surveys. This study was conducted according to the guidelines laid down in the Declaration of Helsinki and all procedures involving research study participants were approved by Institutional Review Boards in the respective countries.

### Estimating coverage of SPPs

Coverage for each SPP was defined as the percentage of households with individuals that reported receipt of government cash or food transfers in the past 12 months. For three SPPs targeting pregnant women (the Janani Surakshya Yojana cash transfer in India, food supplements under India’s Integrated Child Development Services (ICDS), and the Aama Karyakram cash transfer in Nepal), coverage was calculated among women with a pregnancy within the past five years. After reporting coverage of each SPP, we summarized the percentage of households covered by any SPP that provided cash and/or food (four aggregate indicators: any cash, any food, any cash or any food, any cash and any food). For example, if there were four SPPs with cash transfers measured in a given survey, the “any cash” coverage indicator was the percentage of households that received transfers from at least one of those SPPs. If there were four cash SPPs and one food SPP measured in a survey, the “any cash and any food” indicator was the percentage of households covered by at least one cash SPP and the food SPP.

### Estimating co-coverage of SPPs and health/nutrition interventions

Co-coverage was defined as receiving both a SPP transfer and key health/nutrition interventions at three different life stages typically noted as being nutritionally vulnerable periods: during the last pregnancy, after delivery, and during childhood under five years of age. The four health/nutrition interventions considered during pregnancy included 1) at least four antenatal care (ANC) visits, 2) at least 100 iron and folic acid (IFA) tablets consumed, 3) maternal deworming, and 4) two or more tetanus injections. These same four interventions were used for co-coverage of interventions with cash after delivery. For early childhood, the five health/nutrition interventions considered, based on data availability, were: 1) vitamin A supplementation in the past 6 months, 2) deworming in the past 6 months, 3) iron syrup in the past 7 days, 4) growth monitoring, and 5) counseling on nutritional status after growth monitoring. Co-coverage of the SPP and each health/nutrition intervention was estimated, followed by the target scenario of co- coverage of the SPP and all health/nutrition interventions combined. For example, the first co-coverage indicator was the percentage of women with children under five years who received any food transfer during pregnancy and at least four ANC visits during last pregnancy. The second co-coverage indicator was the percentage of women with children under five who received any food transfer during pregnancy plus at least 100 IFA tablets during last pregnancy, and so forth. Similar co-coverage indicators were estimated for food plus deworming and food plus tetanus injections. The fifth and final co-coverage indicator was for an optimal scenario: the percentage of women who received the food transfer plus all four health/nutrition interventions during their pregnancy. The same approach was taken for cash transfers after delivery and food transfers during childhood. Data were not available for cash transfers during pregnancy, food transfers after delivery, or cash transfers during early childhood.

## RESULTS

### Description of survey data on SPPs and sample characteristics

In Bangladesh, coverage data were available for 11 and 6 SPPs, respectively in HIES 2016 and BMICS 2019 (**Table 1**). In India, data were available for 2 SPPs in both NSS 2012 and NFHS 2016. In Nepal, data were available only for a maternal conditional cash transfer program in NDHS 2016 and for 5 types of SPPs in NMICS 2016. No surveys had data on public charity programs. Included programs are listed in Table 1, with additional details for each program provided in **Supplementary Table 1**. The total number of households included in the surveys ranged from 11,040 in NDHS 2016 to 601,509 in NFHS 2016, and 20-40% of households had children under 5 years of age (**Supplementary Table 2**). Supplementary Table 2 also provides additional household characteristics. **Coverage of SPPs and health/nutrition interventions**

**Table 1.** Data availability on social protection programs in population-based surveys in Bangladesh, India, and Nepal^1^.

| Program subcategory | Bangladesh |  |  | India |  |  | Nepal |  |  |
| --- | --- | --- | --- | --- | --- | --- | --- | --- | --- |
|  | Data availability |  | Program name(s) | Data availability |  | Program name(s) | Data availability |  | Program name(s) |
|  | HIES 2016 | BMICS 2019 |  | NSS 2012 | NFHS 2016 |  | NDHS 2016 | NMICS 2019 |  |
| Cash for work | P | P | Work for Money; Test relief; Employment generation program for the poorest | O | O | The Mahatma Gandhi National Rural Employment Guarantee | O | O | Prime Minister Employment program |
| UCT |  |  |  |  |  |  |  |  |  |
| Poverty-targeted cash | P | O | Targeted ultra-poor program | NA | NA | - | O | P | Endangered Indigenous Peoples Allowance or Endangered Ethnicity Grant |
| Allowances |  |  |  |  |  |  |  |  |  |
| Old age | P | P | Old-age Allowance program | O | O | Indira Gandhi National Old Age Pension Scheme | O | P | Old Age Allowance or Senior Citizen's Allowance |
| Widow/Single women | P | P | Husband-Deserted; Widowed and Destitute Women Allowance | O | O | Indira Gandhi National Widow Pension Scheme | O | P | Single Women's Allowance |
| Disabled | P | O | Allowance for Financially Insolvent Persons with Disabilities | O | O | Indira Gandhi National Disability Pension Scheme | O | P | Disability Grant |
| Martyr's<sup>2</sup> | P | P | Allowance for freedom fighters/Shahid family | NA | NA | - | - | - | - |
| Public charity | NA | NA | - | NA | NA | - | NA | NA | - |
| CCT |  |  |  |  |  |  |  |  |  |
| Women/Pregnant women | P | P | Maternity allowance program for the poor lactating mothers | O | P | Janani Suraksha Yojana | P | O | Aama program |
| Students | P | O | Primary Education Stipend program; Secondary Education Sector Investment program; Anand School program | NA | NA | - | O | O | Scholarships program |
| Food transfer | P | P | Vulnerable Group Development/ Vulnerable Group Feeding | P | P | Food supplements to pregnant and lactating women and children under Integrated Child Delivery Services program | O | O | Supplementary food for lactating women and children in food insecure areas |

| Program subcategory | Bangladesh |  |  | India |  |  | Nepal |  |  |
| --- | --- | --- | --- | --- | --- | --- | --- | --- | --- |
|  | Data availability |  | Program name(s) | Data availability |  | Program name(s) | Data availability |  | Program name(s) |
|  | HIES 2016 | BMICS 2019 |  | NSS 2012 | NFHS 2016 |  | NDHS 2016 | NMICS 2019 |  |
| Nutrition program<sup>3</sup> | P | O | Improving Maternal and Child Nutrition; Community Nutrition program | NA | NA | - | O | P | Child (0-5 years) grant |
| School feeding | P | O | School feeding program in the poverty-prone areas | P | O | Mid-Day Meal | O | O | National School Meals program; Food for Education |

<sup>1</sup>For a more detailed description of each program, refer to Supplementary Table 1
<sup>2</sup>Family of a person who died in a war
<sup>3</sup>Any transfer (cash or food) that may be linked to the fulfillment of nutrition practices such as completing growth monitoring
P= Program and data available; O= Program available but data not available; NA=Program does not exist, thus data availability assessment was not applicable
Abbreviations: CCT=conditional cash transfer; HIES=Household Income and Expenditure Survey; MICS=Multiple Indicator Cluster Survey; NDHS=Nepal Demographic and Health Survey; NFHS=National Family Health Survey; NSS=National Sample Survey; UCT=unconditional cash transfer

Coverage of SPPs in Bangladesh and Nepal was less than 20% in nearly all cases and less than 5% in many cases, while about half of households were covered in India (**Table 2**). Between 12% (Bangladesh) and 41% (India) of households received at least one cash transfer program. Coverage of at least one food transfer program was highest in India, where 60% of households received take home rations through the ICDS program. In Bangladesh, only 3% of households received a food transfer through the Vulnerable Group Development program or a school feeding program per HIES 2016 data. In Bangladesh, 18% of households received either cash or food but less than 2% received both cash and food. In India, due to moderate coverage of Janani Surakshya Yojana and of take home rations through ICDS, 16% of households received both cash and food.

**Table 2.** Coverage of individual social protection programs in the past 12 months in Bangladesh, India, and Nepal^1^.

|  | Bangladesh |  | India |  | Nepal |  |
| --- | --- | --- | --- | --- | --- | --- |
|  | HIES | BMICS | NSS | NFHS | NDHS | NMICS |
|  | 2016 | 2019 | 2012<sup>4</sup> | 2016 | 2016 | 2019 |
|  | % | % | % | % | % | % |
| **Social protection programs** |  |  |  |  |  |  |
| Cash for work |  |  |  |  |  |  |
| Employment generation | - | 1.2 | - | - | - | - |
| Gratuitous relief | 1.4 | - | - | - | - | - |
| Test relief | 0.6 |  |  |  |  |  |
| Unconditional cash transfer |  |  |  |  |  |  |
| Old age allowance | 4.1 | - | - | - | - | 12.9 |
| Widow allowance | 1.3 | - | - | - | - | 6.8 |
| Disabled allowance | 0.4 | - | - | - | - | 1.3 |
| Any unconditional allowances<sup>2</sup> | - | 10.2 | - | - | - | - |
| Conditional cash transfer |  |  |  |  |  |  |
| Anand school program | 0.4 |  |  |  |  |  |
| Stipend for primary school students | 6.9 | - | - | - | - | - |
| Stipend for secondary school students | 2.7 | - | - | - | - | - |
| Maternity allowance | - | 0.8 |  |  |  |  |
| Janani Surakshya Yojana<sup>3</sup> | - | - | - | 41.1 | - | - |
| Aama Karyakram<sup>3</sup> | - | - | - | - | 86.4 | - |
| Nutrition program |  |  |  |  |  |  |
| Child support grant | - | - | - | - | - | 2.9 |
| Food support |  |  |  |  |  |  |
| VGD/VGF | 2.5 | 7.4 | - | - | - | - |
| Public distribution system | - | - | 44.1 | - | - | - |
| Food supplement for pregnant women under ICDS<sup>3</sup> | - | - | - | 51.9 | - | - |
| Food supplement for children under ICDS<sup>3</sup> |  |  |  | 51.3 |  |  |
| School feeding | 0.9 | - | 37.9 | - | - | - |
| **Transfer type received (aggregate indicators)** |  |  |  |  |  |  |
| Any cash | 15.9 | 11.6 | - | 41.1 | 86.4 | 22.1 |
| Any food | 3.3 | 7.4 | 51.8 | 59.8 | - | - |
| Any cash or any food | 18.3 | 17.7 | - | 65.9 | - | - |
| Any cash and any food | 1.0 | 1.6 | - | 15.7 | - | - |
| **Nutrition/health interventions** |  |  |  |  |  |  |
| Pregnancy |  |  |  |  |  |  |
| ≥ 4 ANC visits, % | - | - | - | 49.3 | 69.4 | - |
| ≥ 100 IFA, % | - | - | - | 30.3 | 65.4 | - |
| Deworming, % | - | - | - | 18.0 | 70.2 | - |
| Two tetanus injections, % | - | - | - | 83.5 | 66.1 | - |
| Early childhood |  |  |  |  |  |  |
| Vitamin A in past 6m (6-59m), % | - | - | - | 57.8 | 86.0 | - |
| Deworming in past 6m (12-59m), % | - | - | - | 33.0 | 75.8 | - |
| Iron syrup in past 7 days (6-59m), % | - | - | - | 26.0 | 7.7 | - |
| Growth monitored (0-59m), % | - | - | - | 43.3 | 25.3 | - |
| Counseling after growth monitoring (0-59m), % | - | - | - | 27.8 | 12.2 | - |

<sup>1</sup>For a more detailed description of each program, refer to Supplementary Table 1
<sup>2</sup>BMICS 2019 asked about all allowances combined (old age/disabled/widow/freedom fighters/shared families, etc.)
<sup>3</sup>The recall period for these programs was the previous pregnancy (not necessarily the last 12 months).
<sup>4</sup>In India, NSS data is shown for the percentage of households that received rice or wheat in the past 30 days or with children 6-14 years old who received a free school meal in the past 30 days.
*Abbreviations:* HIES=Household Income and Expenditure Survey; ICDS=Integrated Child Development Service; BMICS= Bangladesh Multiple Indicator Cluster Survey; NDHS=Nepal Demographic and Health Survey; NFHS=National Family Health Survey; NMICS= Nepal Multiple Indicator Cluster Survey; NSS=National Sample Survey; VGD=Vulnerable Group Development; VGF=Vulnerable Group Feeding.

Data on health/nutrition interventions were available for India (NFHS 2016) and Nepal (NDHS 2016) but not for Bangladesh surveys. Coverage of interventions during pregnancy was lower in India compared to Nepal, except for tetanus injections. There was large variation in coverage of interventions during early childhood, ranging from 26% (IFA syrup) to 58% (vitamin A supplementation) in India and from 8% (IFA syrup) to 86% (vitamin A supplementation) in Nepal.

### Co-coverage of SPPs and health/nutrition interventions

Among the six surveys, only NFHS 2016 and NDHS 2016 had data on both SPPs and health/nutrition interventions. Although half (52%) of women in India with a birth in the past 5 years received a food transfer during their pregnancy, a take home ration through the ICDS scheme, only 3% received a food transfer plus four health/nutrition interventions: having at least four ANC visits, consuming at least 100 IFA tablets, receiving deworming tablets, and receiving two tetanus toxoid injections (**Figure 1A**).

**Figure 1.**
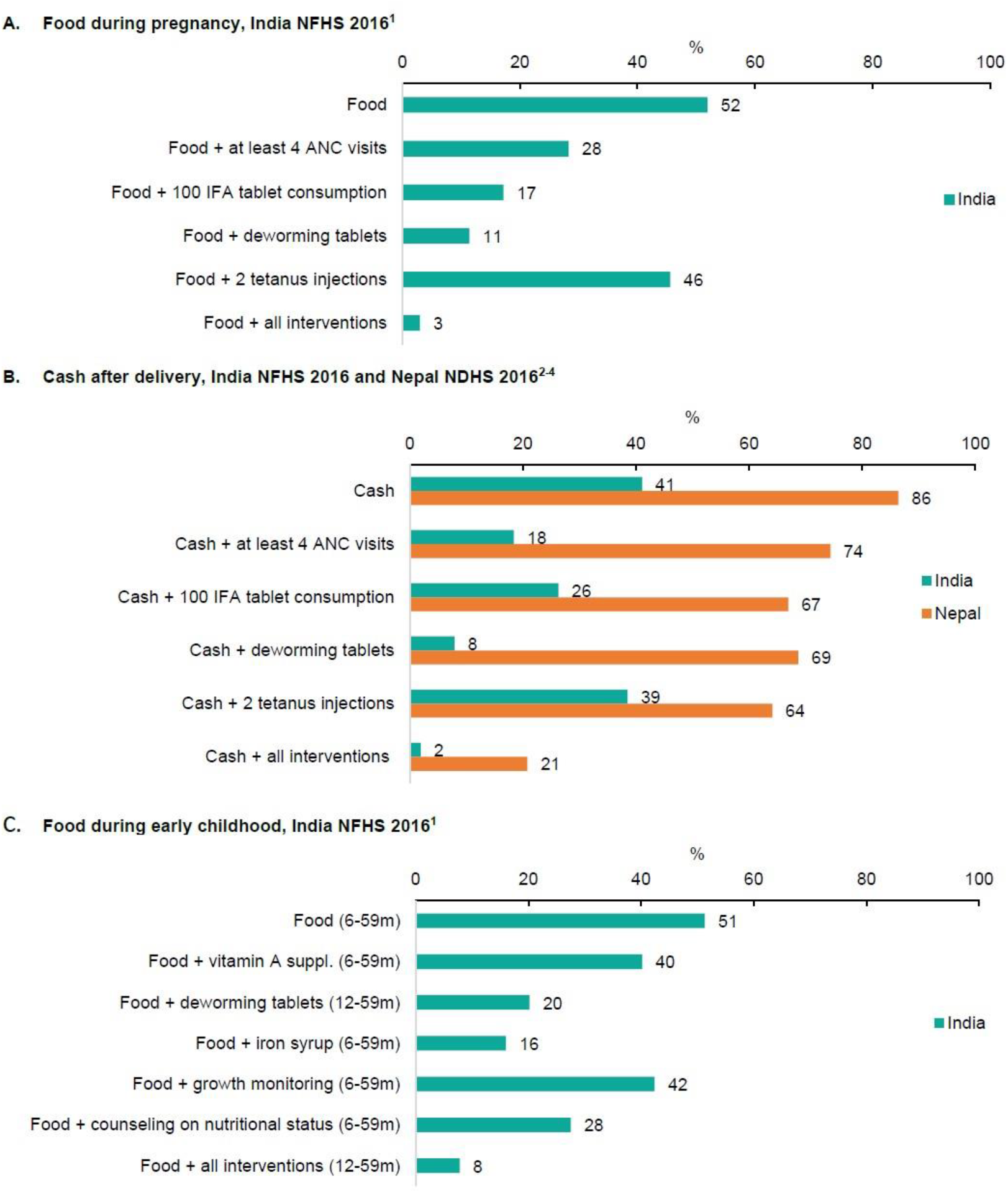
Co-coverage of transfers with other health/nutrition interventions during pregnancy (A), after delivery (B) or in early childhood (C) in India and Nepal. ^1^Food in India’s NFHS 2016 refers to food supplementation during pregnancy and during early childhood under the Integrated Child Development Services program; the denominator for this analysis was women who gave birth in the past 5 years for women and denominators varied by interventions for children according to age group. ^2^Among women who gave birth in a public health facility in the past 5 years. ^3^Cash in India’s NFHS 2016 refers to cash received through Janani Surakshya Yojana. ^4^Cash in Nepal’s NDHS 2016 refers to cash received through Aama Karyakram. Abbreviations: ANC=antenatal care; IFA=iron folic acid; NDHS=Nepal Demographic and Health Survey; NFHS=National Family Health Survey

Similarly, in India and Nepal, respectively, while 41% and 86% of women received cash through perinatal cash transfer programs, only 2% and 21% of women received cash plus all four health/nutrition interventions (**Figure 1B**). Co-coverage of India’s ICDS food transfer and health/nutrition interventions during early childhood followed the same pattern, with 51% of children getting the transfer but only 8% of children getting the transfer plus all five health/nutrition interventions (**Figure 1C**).

## DISCUSSION

While many SPPs exist in South Asia, measurement of whether households and/or target populations receive benefits and related program features in population-based surveys remains poor (15). While coverage on health/nutrition interventions exist, few surveys measure coverage of both SPPs and health/nutrition interventions simultaneously or if SPP benefits include linked health/nutrition interventions. Using available data from South Asia, our study found low coverage of most SPPs and a missed opportunity to reach households with both SPP and health/nutrition interventions.

Many examples of large scale SPPs exist in South Asia, but they are infrequently measured. Although social protection systems and, thus, SPPs differ across countries, certain SPPs that address underlying determinants of malnutrition, such as school feeding programs or perinatal cash transfers, exist across contexts in South Asia. Therefore, measuring both common SPP elements and generic features proven to improve nutrition outcomes through a standard set of questions in household surveys would enable cross-country comparisons of SPP coverage. Questions that ask about additional details on the transfers, (e.g. quantity of cash received or school meal quality), would allow for further program refinement and explorations of how program quality relates to measured nutrition outcomes. In the absence of such data on the potentially nutrition-sensitive features of SPPs, co-coverage is a useful approach to understand if households are receiving multiple interventions, but the scope of co-coverage estimation remains limited due to few surveys measuring receipt of both SPPs and health/nutrition interventions.

While these six surveys are not an exhaustive list of surveys with SPP measurement in South Asia, we used readily accessible datasets that allowed us to explore the concept of SPP and health/nutrition intervention co-coverage. Our findings are generally in agreement with those from a phone survey in India conducted during the COVID-19 pandemic that included detailed questions on receipt of SPPs and health/nutrition interventions by women (13). Co-coverage estimates in the phone survey were higher than what we found in the current study, likely because the phone survey was more recent (2022 compared to 2016 for NFHS), thus existing programs had wider reach and there were new programs; and the phone survey included a broader set of SPPs and health/nutrition interventions compared to NFHS. SPP coverage estimates from nationally representative surveys such as DHS and MICS may not account for all existing SPPs within a country. Notably, Living Standards Measurement Study surveys typically measure SPP coverage but do not measure receipt of health/nutrition interventions. Many non-governmental organizations may provide social protection services in focused geographies, which are often not tracked in large scale surveys. In any given survey, there is a limitation to the amount of information that can be collected, thus multiple data sources should be utilized when possible. Some countries have administrative data systems that track social assistance disbursed to beneficiaries. The quality and frequency of administrative data varies, and this data source is often not available to the public. Future research should triangulate information from multiple data sources to more closely examine SPP coverage and co-coverage of multisectoral interventions.

SPPs or health/nutrition interventions in isolation have limited scope to respond to the broad range of shocks and challenges that households face during critical periods of growth and development. Ensuring that vulnerable households are reached by complementary interventions that provide both social protection transfers and health/nutrition interventions to address underlying and intermediate needs is critical for optimal maternal, infant and young child wellbeing. Measurement of SPPs and health/nutrition interventions in population-based surveys requires further investment.

## Data Availability

All data used in the study are publicly available.
Bangladesh Household Income and Expenditure Survey (HIES) 2016 is available at: https://catalog.ihsn.org/index.php/catalog/7399/related-materials
Bangladesh Multiple Indicator Cluster Survey (BMICS) 2019 is available at: https://mics.unicef.org/surveys
India National Family Health Survey (NFHS) 2016 is available at: https://dhsprogram.com/data/
India National Sample Survey (NSS) 2012 is available at:
https://microdata.gov.in/nada43/index.php/catalog/127/study-description
Nepal Demographic and Health Survey (NDHS) 2016 is available at: https://dhsprogram.com/data/
Nepal Multiple Indicator Cluster Survey (NMICS) 2019 is available at: https://mics.unicef.org/surveys

## Acknowledgements

We appreciate comments from Swetha Manohar, a Research Fellow at the International Food Policy Research Institute, on a draft of this manuscript.

## Abbreviations

ANC: antenatal care
BMICS: Bangladesh Multiple Indicator Cluster Survey
CCT: conditional cash transfer
DHS: Demographic and Health Survey
HIES: Household Income and Expenditure Survey
ICDS: Integrated Child Development Services
IFA: iron folic acid
NDHS: Nepal Demographic and Health Survey
NFHS: National Family Health Survey
NMICS: Nepal Multiple Indicator Cluster Survey
NSS: National Sample Survey
SPP: social protection program
UCT: unconditional cash transfer
VGD/VGF: vulnerable group development/vulnerable group feeding

## REFERENCES

1. FAO, IFAD, UNICEF, WFP, WHO. The State of Food Security and Nutrition in the World 2023. The State of Food Security and Nutrition in the World 2023. FAO; IFAD; UNICEF; WFP; WHO; 2023 Jul.

2. Keats EC, Das JK, Salam RA, Lassi ZS, Imdad A, Black RE, et al. Effective interventions to address maternal and child malnutrition : an update of the evidence. Lancet Child Adolesc Health [Internet]. 2021;4642(20):1–18. Available from: 10.1016/S2352-4642(20)30274-1

3. Bhutta ZA, Das JK, Rizvi A, Gaffey MF, Walker N, Horton S, et al. Evidence-based interventions for improvement of maternal and child nutrition: What can be done and at what cost? The Lancet. 2013;382(9890):452–77.

4. Gentilini U. Social Protection, Food Security and Nutrition An Update of Concepts, Evidence and Select Practices in South Asia and Beyond. 2022.

5. Ruel MT, Alderman H. Nutrition-sensitive interventions and programmes: How can they help to accelerate progress in improving maternal and child nutrition? The Lancet. 2013;382(9891):536–51.

6. Ruel MT, Quisumbing AR, Balagamwala M. Nutrition-sensitive agriculture: What have we learned so far? Vol. 17, Global Food Security. Elsevier B.V.; 2018. p. 128–53.

7. Ahmed A, Hoddinott J, Roy S. Food transfers, cash transfers, behavior change communication and child nutrition. Evidence from Bangladesh. 2019. Report No.: 01868.

8. Scott S, Neupane S, Alderman H, Kim S, Parvin A, Rasheed S, et al. Do social protection programs in South Asia have the potential to be nutrition-sensitive? Insights from Bangladesh, India, Nepal, and Pakistan. 2023.

9. Neupane S, Scott S, Jangid M, Shapleigh S, Kim S, Akseer N, et al. Data Availability on Nutrition Sensitive Social Protection Programs (NSSPPs) Across Population-based Surveys in South Asia. 2022.

10. Menon P, Avula R, Pandey S, Scott S, Kumar A. Rethinking Effective Nutrition Convergence An Analysis of Intervention Co-coverage Data. Econ Polit Wkly. 2019;54(24).

11. Nguyen PH, Singh N, Scott S, Neupane S, Jangid M, Walia M, et al. Unequal coverage of nutrition and health interventions for women and children in seven countries. Bull World Health Organ [Internet]. 2022 Jan 1 [cited 2023 Oct 17];100(1):20. Available from: /pmc/articles/PMC8722629/

12. Victora CG, Fenn B, Bryce J, Kirkwood BR. Co-coverage of preventive interventions and implications for child-survival strategies: Evidence from national surveys. Lancet [Internet]. 2005 Oct 22 [cited 2023 Nov 9];366(9495):1460–6. Available from: http://www.thelancet.com/article/S014067360567599X/fulltext

13. Nguyen PH, Avula R, Neupane S, Akseer N, Heidkamp R. Identifying measures for coverage of nutrition-sensitive social protection programs: Learnings from India. Matern Child Nutr. 2024 Jun 12;

14. World Bank. ASPIRE program classification [Internet]. [cited 2023 Nov 9]. Available from: https://thedocs.worldbank.org/en/doc/05c7cffd57ec07f5a3fc8d598ca4941d-0380052021/original/ASPIRE-program-classification.pdf

15. Gillespie S, Menon P, Heidkamp R, Piwoz E, Rawat R, Munos M, et al. Measuring the coverage of nutrition interventions along the continuum of care: time to act at scale. BMJ Glob Health [Internet]. 2019 May 1 [cited 2024 Sep 3];4(Suppl 4):e001290. Available from: https://gh.bmj.com/content/4/Suppl_4/e001290

