## Supplementary Table for "Co-coverage of social protection programs and maternal and child nutrition interventions in Bangladesh, India and Nepal"

**Supplementary Table 1. Nutrition sensitive social protection programs in Bangladesh, India and Nepal**

| Country | | Name of the Scheme | Eligibility/conditionality | Entitled benefits | Geographic coverage |
|---|---|---|---|---|---|
| Bangladesh | Cash/food for work | Food for work, Test relief, Employment generation program for the poorest | Eligible households should have less than 0.5 acre of land, no productive assets, and less than BDT4,000 (approximately USD50) as annual income; the head of household should also work as a day labor | 80 days of guaranteed employment per year | Nationwide, with priority given to the poorest areas |
| | Allowances | | | | |
| | Old age | Old-age Allowance Program | The age of eligibility is 65 for men and 62 for women. Beneficiaries' annual income must be below BDT10,000 | BDT 500 per month | Nationwide |
| | Disabled/ Marty's | Allowance for Financially Insolvent Persons with Disabilities | Permanent residents living with a hearing, visual, speech, intellectual, and/or physical impairment, as defined by the Bangladesh Disability Welfare Act (2001), who have an annual income lower than BDT36,000 and are older than 6 years of age.<br>Food support for shaheed family and injured freedom fighters | BDT 700 per month | Nationwide |
| | Widow/Single women | Husband-Deserted, Widowed and Destitute Women Allowance, HWDWA | Elderly, People with Disability, Women (widows and husband-deserted women), and Mothers (with more than one child) | BDT 500 per month | Nationwide |
| | Conditional cash transfer | | | | |
| | Women | Mother and Child benefit prog. | Working lactating women | BDT 800 per month until the child is 3 years old | 64 districts |
| | Students | Primary Education Stipend Program, PESP | All primary school children enrolled in participating primary schools | BDT 100 for 1 student BDT 200 for 2 students BDT 250 for 3 students BDT 300 for 4 students in schools that have been expanded up to grades VI–VIII receive BDT125 per student per month Pre-primary students are eligible to receive BDT 50 per month | Nationwide |
| | | Secondary Education Sector Investment Program (SESIP) | School-based committees apply a 'pro-poor methodology' in selecting the poorest students to participate in the program | The yearly stipend rate varies according to the students' grades: Class 6 and class 7 – BDT1380 Class 8 – BDT 1680 Class 9 – BDT 2280 Class 10 – BDT 3510 | Schools and Madrasahas of 54 upazilas under 17 districts |
| | | Anand school program | | | |
| | Food transfer | Public Food Distribution System (PFDS) | Food assistance programs typically target poor people (whose poverty and vulnerability status should be assessed based on land ownership, annual income, and/or occupation) and/or disaster-affected populations. The Vulnerable | Open Market Sales (OMS) – Rates of BDT 30 (USD0.36) per kilogram of rice and BDT 17 (USD0.2) per kilogram of wheat flour for low-income families (around 5 kg of rice per day per family | Nationwide |

**Supplementary Table 1. Nutrition sensitive social protection programs in Bangladesh, India and Nepal**

| Country | | Name of the Scheme | Eligibility/conditionality | Entitled benefits | Geographic coverage |
|---|---|---|---|---|---|
| | | | Group Development (VGD) targets female-headed households whose head is able to work | was set to be distributed until the Boro harvest in April 2018). Food for Work (FfW) and Test Relief (TR) – 8 kg of rice per work day. Work for Money (WfM) – 8 kg of rice/wheat or money equivalent to 7 hours of work Vulnerable Group Development (VGD) – 30 kg of rice per month per household and social support services (including life skills and income-generating skills training, savings and access to credit) for a cycle of twenty-four months. Vulnerable Group Feeding (VGF) - 10-30 kg of rice/wheat per month. Gratuitous Relief (GR) – The average amount of rice given out by the Gratuitous Relief program is 15.7 kilograms per beneficiary. This program also offers other cash or in-kind transfers (e.g. blankets) to meet the needs of disaster-affected people. Food Assistance for the Chittagong Hill Tracts Region – 3.5 kg per day per person (data from 2009). | |
| | School feeding | School Feeding Program in the Poverty-prone Areas | Beneficiaries must be enrolled in primary schools. | One 75-gram packet of fortified biscuits. WFP is running a mid-day meal pilot scheme with cooked food in all primary schools of Bamna Upazila in Barguna district, as well as in the schools of two Unions of Islampur upazila of Jamalpur district | WFP prioritized the poorest regions, with the worst performance in primary education indicators. |
| India | Cash/food for work | Mahatma Gandhi National Rural Employment Guarantee (MGNREG) program | Below poverty line people | At least 100 days of guaranteed wage employment in a financial year to every rural household whose adult members volunteer to do unskilled manual work. | Nationwide |
| | Allowances | | | | |
| | Old age | Indira Gandhi National Old Age Pension Scheme | Below Poverty Line People; 60 years and above | INR 200 per month; INR 500 for the elderly above 80 years of age | Nationwide |
| | Disabled/Marty's | Indira Gandhi National Disability Pension Scheme | Below Poverty Line People; 18 years and above with a disability level of 80 per cent | INR300 per month; INR 500 for the elderly above 80 years of age | Nationwide |
| | Widow/Single women | Indira Gandhi National Widow Pension Scheme | Below Poverty Line People; 40 years and above | INR 300 per month; INR 500 for the elderly above 80 years of age | Nationwide |

**Supplementary Table 1. Nutrition sensitive social protection programs in Bangladesh, India and Nepal**

| Country | | Name of the Scheme | Eligibility/conditionality | Entitled benefits | Geographic coverage |
|---|---|---|---|---|---|
| | Conditional cash transfer | | | | |
| | Women | Pradhan Mantri Matritva Vandana Yojana | Early registration of pregnancy, one antenatal care visit, child registration and vaccination/immunization. All first-time pregnant women and lactating mothers, except those regularly employed by the public sector or who receive similar benefits. | The total of benefits adds up INR5000, which is paid in three installments as listed below: INR1000 for the early registration of pregnancy at an Anganwadi Centre (AWC)– approved health facility INR2000 after six months of pregnancy if the beneficiary attends one antenatal check-up session INR2000 after the child is registered and vaccinated. | Nationwide |
| | | Janani Suraksha Yojana | Beneficiaries are required to give birth in a health facility. In low-performing states, the program covers all women delivering in public health centers regardless of any vulnerability criteria, as well as those women classified as 'below the poverty line' (BPL) and members of a Scheduled Caste or Scheduled Tribe delivering in accredited private institutions. However, in states that already have higher institutional delivery rates, the program is reserved for those considered BPL or Scheduled Castes or Tribes. | For LPS in rural areas: mothers' package of INR1400 and ASHA package of INR600 For LPS in urban areas: mothers' package of INR 1000 and ASHA package of INR400 For HPS in rural areas: mothers' package of INR700 and ASHA package of INR600 For HPS in urban areas: mothers' package of INR600 and ASHA's package of INR400 Women below the poverty line who prefer to deliver at home are also entitled to INR500 ASHAS receive the abovementioned values per pregnant women they enable having prenatal monitoring and institutional delivery | Nationwide |
| | Food transfer | Food transfer to pregnant and lactating women and children under Integrated Child Development Service (ICDS) | All pregnant women and children under 36 months | Pregnant and lactating women receive food supplement of 600 calories of energy and 18-20 grams of Protein per day in the form of Micronutrient Fortified Food and/or energy dense food as Take-Home Ration from Anganwadi Centers. | Nationwide |
| | | Public distribution system | All households | Under the program food rations are available at subsidized rates through Government sponsored shops. Each family below the poverty line is eligible for 35 kg of rice or wheat every month, while a household above the poverty line is entitled to 15 kg of food grain on a monthly basis | Nationwide |
| | School feeding | Mid-Day Meal | Children must be enrolled in public schools (government, government-aided, local body, EGS and AIE Centers, Madrasas and Maqtabs supported under Sarva Shiksha Abhiyan and | Students receive a meal containing 100 grams of food grains, 20 grams of dal, 50 grams of vegetables, and 5 grams of oil and fat (450 calories in total) at the primary stage and 150 grams of food | Nationwide |

**Supplementary Table 1. Nutrition sensitive social protection programs in Bangladesh, India and Nepal**

| Country | | Name of the Scheme | Eligibility/conditionality | Entitled benefits | Geographic coverage |
|---|---|---|---|---|---|
| | | | NCLP Schools run by the Ministry of Labor), studying in primary or upper primary classes | grains, 30 grams of dal, 75 of vegetables, and 7.5 grams of oil and fat (700 calories in total) at the upper primary stage per school day (314 days) | |
| Nepal | Cash/food for work | Prime minister employment program | Any household in Karnali with an unemployed member. The sole enrolment limitation is that those who volunteer for work are of the legal age to do so. | 100 days of guaranteed employment per year | Karnali province |
| | Unconditional cash transfer | | | | |
| | Poverty-targeted cash | Endangered Indigenous Peoples Allowance or Endangered Ethnicity Grant | Beneficiaries are members of the ten ethnic groups which are officially recognized as being endangered (i.e: Bankariya, Hayu, Kisan, Kusbadiya, Kusunda, Lepcha, Meche, Raji, Raute, Surel) | NPR 3,990 per month | Nationwide |
| | Allowances | | | | |
| | Old age | Old Age Allowance (OAA) or Senior Citizen's Allowance | Recipients must be at least 70 years old. Dalits and the residents of the Karnali region must be at least 60 years old to receive the benefit | NPR 2,660-4,000 per month | Nationwide |
| | Disabled/Marty's | Disability Grant | People living with disabilities over the age of 16 qualify for a disability identity card. All those who are fully disabled are eligible to receive the benefit; district-wise quotas apply to the partially disabled | NPR 2,128-3,990 per month | Nationwide |
| | Widow/Single women | Single Women's Allowance | Widows of all ages and single women aged 60 or older are eligible for the grant | NPR 2,660 per month | Nationwide |
| | Conditional cash transfer | | | | |
| | Women | Aama Karyakram | Women receive the benefits upon completion of four antenatal care visits and upon having an institutional delivery in public facilities | NPR 1,000-3,000 for delivery and NPR 400 for completing 4 antenatal care visits | Nationwide |
| | Students | Scholarships | Dalit children, girls, people with disabilities, 'endangered' and marginalized groups, conflict-affected children and children of martyrs are eligible for scholarship program; Scholarship recipient should have 70% attendance at the school | Benefits across the different scholarships range from NPR350 to NPR24,000 per year, depending on the type of scholarship, region and level of education. Some scholarships provide money to students and schools, while others just to the students | Nationwide, with priority given to mountain and hill areas |
| | Food transfer | Mother Child Health and Nutrition program | Pregnant women and children | Pregnant women and children under five years old receive 3 kg of supplementary food every month | Karnali province (6 districts) |
| | Nutrition program | Child (0-5 years) grant | All children in 25 districts are eligible for the grant, from birth until the age of 5. In other regions of the country, all poor dalit children under the age of 5 are eligible. In all cases, birth registration is | NPR 532 per month | Planned to be nationwide, currently only providing universal coverage in |

**Supplementary Table 1. Nutrition sensitive social protection programs in Bangladesh, India and Nepal**

| Country | | Name of the Scheme | Eligibility/conditionality | Entitled benefits | Geographic coverage |
|---|---|---|---|---|---|
| | | | required and a maximum of two children per family may participate in the program | | the Karnali region, whereas in other regions it is limited to Dalit children (a more vulnerable group) |
| | School feeding | National School Meals Program, NSMP, and Food for Education | Children should be enrolled in primary education in the selected districts | Meals provided with the support of WFP consist of 110 grams of corn and soya blend (CSB), flour (90g), sugar (10g) and ghee/oil (10g), amounting to 470 calories (kcal) | 29 out of 75 districts |

Source: www.socialprotection.org

**Supplementary Table 2. Characteristics of households included in surveys from Bangladesh, India and Nepal**

| | Bangladesh | | India | | Nepal | |
|---|---|---|---|---|---|---|
| | HIES 2016 | BMICS 2019 | NSS 2012 | NFHS 2016 | NDHS 2016 | NMICS 2019 |
| Household characteristics | | | | | | |
| Households, n | 46,083 | 61,242 | 101,662 | 601,509 | 11,040 | 12,800 |
| Households with <5y children, % | 15,611 | 23,099 | 27,022 | 180,227 | 2,422 | 5,322 |
| Household head is female, % | 12.8 | 12.7 | 11.6 | 14.6 | 31.3 | 28.0 |
| Rural households, % | 70.0 | 77.9 | 58.7 | 65.1 | 37.2 | 32.7 |
| Agriculture land ownership, % | 33.1 | 37.7 | 40.2 | 38.9 | 77.5 | 77.1 |
| Average land holding, acres | 1.4 | 1.1 | 3.5 | - | - | 1.2 |
| Below poverty line[1], % | 12.9 | - | 21.9 | - | - | - |
| Family size, no. persons | 4.0 | 4.3 | 4.6 | 4.7 | 5.4 | 4.3 |
| Wealth index[2] | - | 0.14 | - | 0.23 | 0.32 | 0.23 |
| Child nutrition outcomes | | | | | | |
| Stunting[3], % | - | 28.0 | - | 38.4 | 35.8 | 31.5 |
| Wasting[4], % | - | 8.8 | - | 21.0 | 9.7 | 12.0 |

*Abbreviations*: HIES=Household Income and Expenditure Survey; MICS=Multiple Indicator Cluster Survey; NSS=National Sample Survey; NDHS=Nepal Demographic and Health Survey; NFHS=National Family Health Survey

[1]Households whose total spending on food items over a specific duration is equal to or less than the amount required to purchase food basket sufficient to supply a specific number of calories per person per day.

[2]Derived by running principal component analysis on household assets and housing characteristics; not comparable across countries.

[3]Children (0-59 months) whose height-for-age z-score is more than 2SD below the median compared to the WHO Child Growth Standards

[4]Children (0-59 months) whose weight-for-height z-score is more than 2SD below the median compared to the WHO Child Growth Standards
